# Improving the detection of clinically significant steatotic liver disease using a machine learning algorithm in a real-world primary care population

**DOI:** 10.64898/2026.03.04.26347631

**Authors:** Huw Purssell, Lucy Bennett, Mohamed Mostafa, Stephanie Landi, Christopher Mysko, Richard Hammersley, Manish Patel, Jennifer Scott, Oliver Street, Karen Piper Hanley, The ID LIVER Consortium, Neil A Hanley, Joanne R Morling, Indra Neil Guha, Varinder S Athwal

## Abstract

**Background and aims:** Population screening for liver disease in high-risk groups is recommended. Community diagnosis of liver disease is a challenge due to the asymptomatic nature of disease until very advanced stages. Moreover, regional variation in testing availability can result in people with clinically significant liver disease being missed. Machine learning (ML) has been proposed as a method to reduce diagnostic error and automate screening. We present a novel machine learning derived algorithm (ID LIVER-ML) designed to predict the risk of clinically significant liver disease in a high-risk community population to identify those needing further investigations or specialist referral.

**Methods:** Using data from 2039 patients recruited to two UK cohorts, we created a parsimonious model using investigations that would be available in primary care using liver stiffness measurement as reference standard. The performance of ID LIVER-ML was compared against FIB-4 score in a second unseen hold out cohort (n=327).

**Results:** ID LIVER-ML performed well at identifying patients at risk of clinically significant liver fibrosis (sensitivity 0.90, Specificity 0.43, PPV 0.54, NPV 0.86, AUC 0.83) and outperformed conventional risk scoring systems (FIB-4: AUC 0.65; NAFLD Fibrosis Score: AUC 0.66; APRI: AUC 0.53; BARD: AUC 0.58).

**Conclusion:** Machine learning derived algorithms can help screen high risk populations in a community setting for liver fibrosis.

**ClinicalTrials.gov ID:** NCT04666402

**Impact and Implications:** The prevalence of steatotic liver disease is rising globally and is an increasingly significant challenge for healthcare systems. Existing risk stratification scores are not validated in a real-world cohort where patients have risk factors for multiple aetiologies of liver disease. Our work shows that a machine learning model can predict the risk of clinically significant liver disease using routine primary care data, better than existing non-invasive risk stratification tools in a real-world cohort. This highlights a potential role for machine learning in the automation of fibrosis risk assessment in primary care.

**Highlights:**

- Machine learning derived algorithms can predict the risk of clinically significant liver disease in an at risk community population with a mixed aetiology of liver diseases.
- The performance of the ML algorithm (ID LIVER-ML) is not affected by metabolic, alcohol, or mixed aetiologies.
- ID LIVER-ML outperforms traditional risk stratification scoring systems such as FIB-4 and NAFLD fibrosis scores.
- Compared to the FIB-4 score, the use of Machine Learning can reduce the need for secondary care investigations by 59%.

## Introduction

The prevalence of steatotic liver disease is rising globally and is an increasing challenge for healthcare systems [1]. Currently, the majority of patients are first diagnosed with liver disease following presentation to hospital with features of decompensated liver cirrhosis [2]. With an increasing incidence of obesity, Type 2 diabetes mellitus (T2DM), metabolic syndrome and increased alcohol consumption an exponential rise in lifestyle related chronic liver disease (CLD) is forecast [3]. Clinically significant liver disease is defined as Metavir Fibrosis stage F2 or above [4]. Identification of patients with clinically significant liver disease as a result of steatotic liver disease may allow intervention to halt, or even reverse, disease progression. The diagnostic challenges of identifying significant liver disease have been well documented and include the poor sensitivity of routine liver blood tests [5, 6] and the cost and widespread availability of specialist fibrosis tests. Whilst the application of cost-effective solutions such as FIB-4 have been advocated [7, 8], there is increasing evidence of limitations including missed disease [6, 9].

Integrated Diagnostics for Early Detection of Liver Disease (ID-LIVER) is a partnership of academics, NHS clinicians and industry aiming to address this challenge by developing population level diagnostic tools for liver disease using novel technologies [10].

Machine Learning (ML) has the potential to offer cost-effective solutions in resource constrained systems. It is set to play a large part in modern medicine due to many reasons, of which limited resources and diagnostic error of humans are two major drivers for change [11]. We have previously described that ML shows promising performance at predicting the risk of liver fibrosis using a single cohort [12]. In this paper, the team describe the creation of a screening tool (ID LIVER-ML) using two separate cohorts which could be used in clinical practice to screen a high-risk population for the presence of clinically significant liver disease. Developing on our previously described methodology with iteratively advanced ML technology to create a diagnostic algorithm fit for clinical practice [12].

## Methods

### Patient selection

For this study, we have used data from two independent cohorts from two large UK cities with high rates of liver disease. In both cohorts, patients were aged over 18 and gave written informed consent for initial data collection and use of data for ongoing research purposes.

Patients already known to have CLD or be under the care of the hospital liver team were excluded. Vector-controlled transient elastography (TE) was performed by trained operators with a liver stiffness measurement (LSM) threshold of 8.0 kPa applied to identify a risk of clinically significant liver disease. After identification patients were invited to a single clinic appointment where a clinical history, blood tests for a full liver aetiological screen and TE (Fibroscan, Echosens, Paris) were performed

### Scarred Liver Project

Patients were recruited prospectively in Nottingham as part of the Scarred Liver Project (SLP) from 2012 to 2016 [6, 13]. The project screened over 25,000 patients from 5 different general practices (GPs) for risk factors for liver disease using READ codes for type 2 diabetes (T2DM), obesity (Body Mass Index (BMI) >30m^2^), abnormal ALT or a history of above recommended alcohol consumption. 1453 patients were recruited.

### ID LIVER cohort

A separate prospective cohort was collected as part of this study in a different region of high disease prevalence. Patients were prospectively recruited to the ID LIVER project in Manchester, UK (https://sites.manchester.ac.uk/id-liver/). Ethical approval was granted as part of the ID LIVER project by REC North of Scotland (REC reference: 20/NS/0055).

Patients with risk factors for liver disease were either identified by primary care physicians or identified pro-actively through interrogation of GP practice records using the Feasibility & Recruitment System for Improving Trial Efficiency (FARSITE) tool developed by North West e-Health, UK. FARSITE used SNOMED codes to identify patients with the following risk factors: T2DM, obesity (BMI>30kg/m^2^), a history of/current harmful alcohol consumption (>50 units alcohol/week in males and >35 units alcohol/week in females), a serum Alanine aminotransferase (ALT) >80 IU/L or a coding of steatosis of the liver. Records were screened in November 2020. Only patients with a history of a BMI >30kg/m^2^, harmful alcohol consumption or abnormal ALT in the preceding 24 months were included.

Overall, 1108 patients were recruited in Manchester as part of this study which was split into two cohorts as outlined in Figure 1.

**Figure 1:**
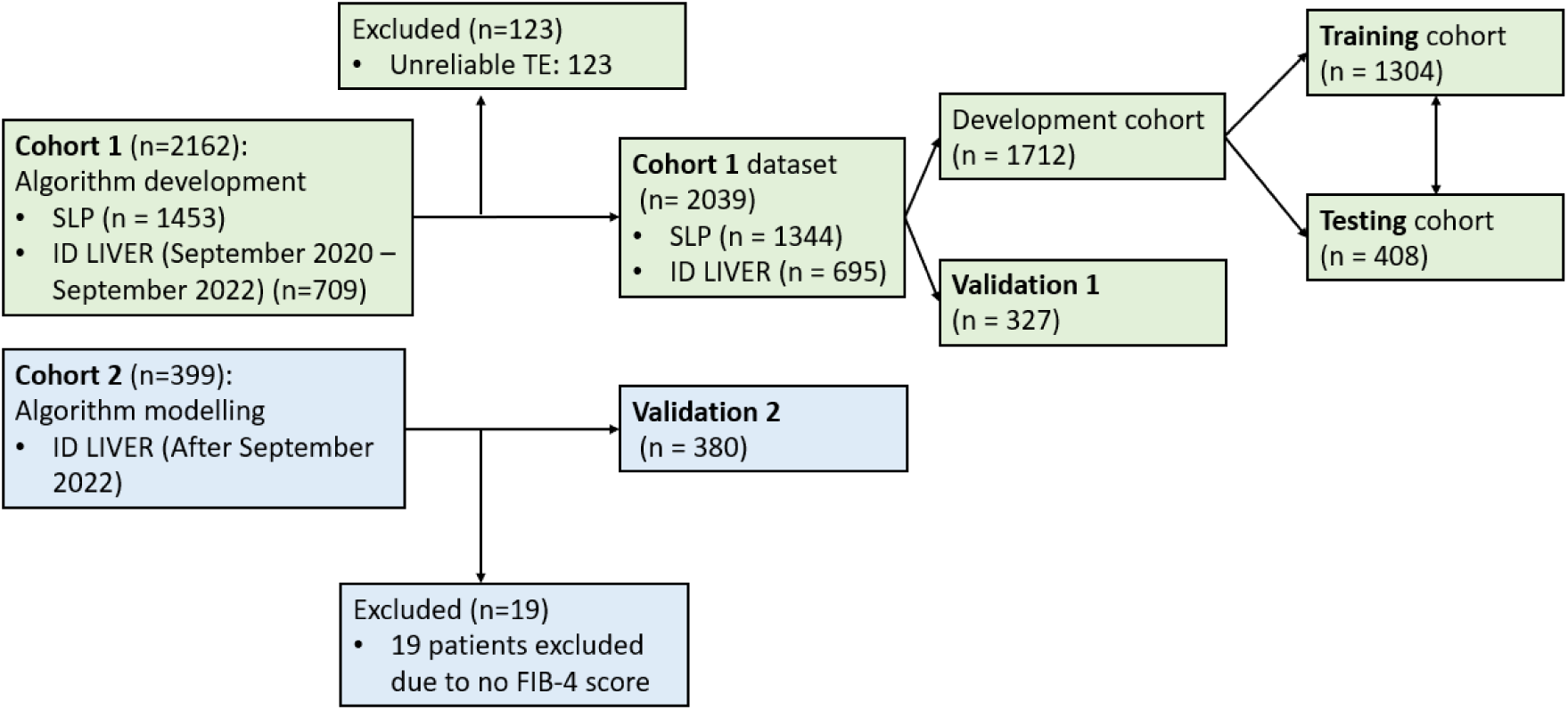
Flowchart showing the cohort data segmentation.

### Cohort selection

We combined two separate datasets (SLP and ID LIVER dataset prior to October 2022) to create Cohort 1 (Figure 1). Merging the datasets not only increased the population available but also added heterogeneity, improving both algorithm power and applicability. The cohort of 2162 patients (1453 patients from the SLP and 709 patients from the ID LIVER project) was used in the initial development, training and validation of ID LIVER-ML.

A separate, prospective cohort of 399 patients recruited to the ID LIVER project between October 2022 to May 2023 was used as an unseen cohort (Cohort 2) for a) a second validation dataset (Validation 2) and b) assess a real-world comparison between ID LIVER-ML and FIB-4 score. Figure 1 shows the cohort split. All datasets were curated using Microsoft Access 2016 and Microsoft Excel.

### ML Algorithm Development and head-to-head comparison with conventional risk stratifying scoring systems

#### Variable selection and Dataset split

67 common socio-demographic and biochemical variables collected in both the ID LIVER and SLP patient populations were included in analysis. Non-synchronous data variables in either dataset were not included for ML algorithm training. Common variables included: age, gender, BMI, comorbidities, medication and alcohol consumption. Serum measurements of bilirubin, ALT, aspartate aminotransferase (AST), alkaline phosphatase (ALP), albumin, sodium, creatinine, haemoglobin, mean corpuscular volume, platelets, fasting cholesterol and lipids and HbA1c were also included for analysis. Natural language processing (NLP) was used to extract open-text variables and automatic application of the International Classification of Diseases 11^th^ Revision was carried out. This was validated by clinicians (HP & LB). Medications were compared with a pre-formulated list based on the British National Formulary and if prescribed, a positive classification was assigned. LSMs were dichotomised using a threshold of 8.0 kPa to determine the presence of clinically significant liver disease. Patients with a TE IQR/Median measurement >30% were removed from the dataset.

Cohort 1 (n = 2039 patients) was split randomly; 80% into datasets for training (n = 1304) and testing (n = 408) and 20% as an unseen hold-out dataset for Validation 1 (n = 327) (Figure 1). The training subset was used to develop ID LIVER-ML, while the testing subset (n = 408) was used to evaluate its performance. The Validation 1 subset was then used to fine-tune the ID LIVER-ML’s hyperparameters, to avoid any level of bias or overfitting. K-fold cross-validation was implemented by dividing the data into K equally sized subsets and using each subset as the testing set while the remaining data is used for training. The process was then repeated K times, with each subset being used as the testing set once. This ensured that ID LIVER-ML was evaluated on unseen data, and for each analysis the results were averaged, providing a more reliable estimate of the algorithm’s performance. The split datasets were matched for the prevalence of risk factors, age, gender and ethnicity.

#### ID LIVER-ML development pipeline Preprocessing

Figure 2 shows the ID LIVER-ML development pipeline.

**Figure 2:**
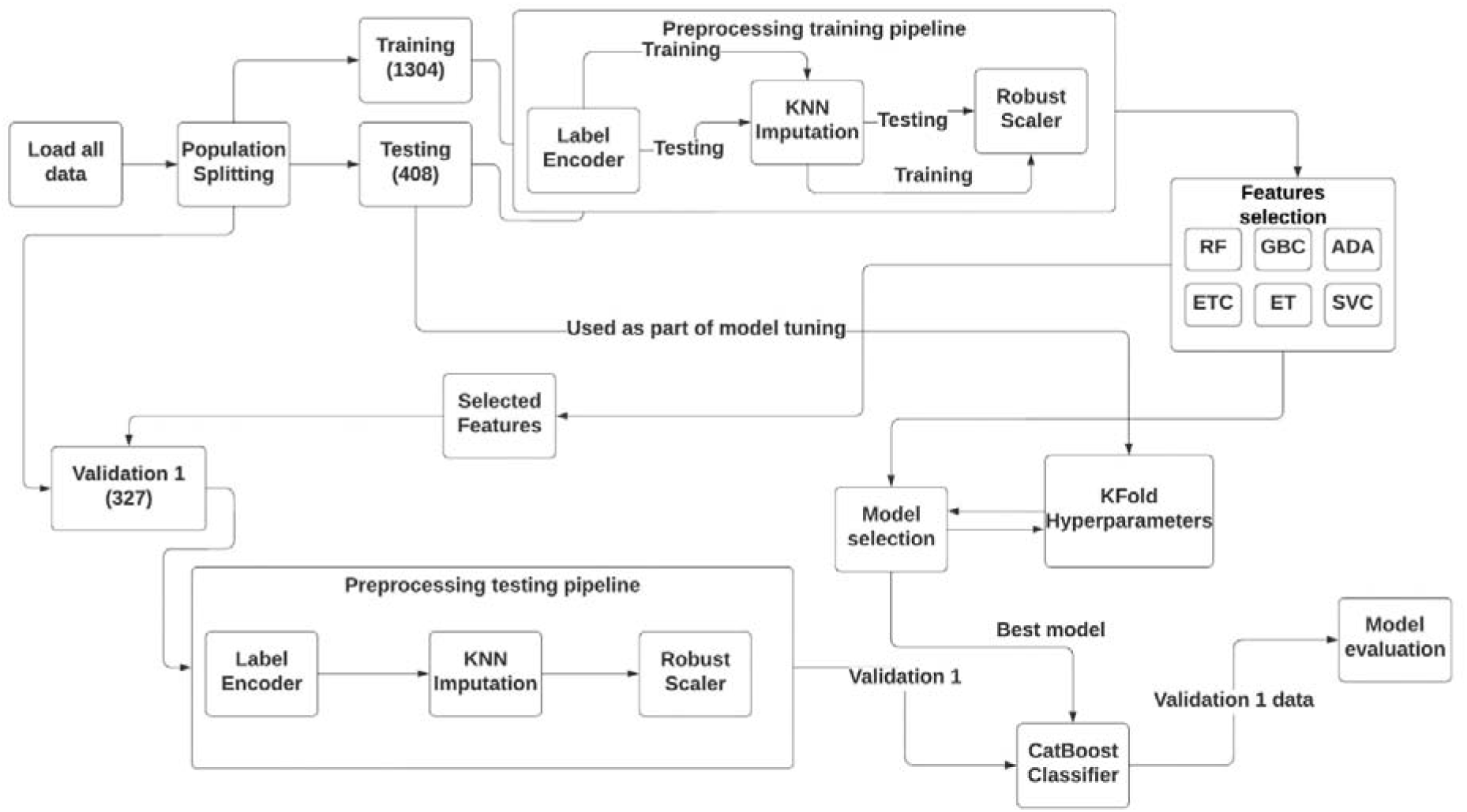
Process map demonstrating the major steps in development of ID LIVER-ML. KNN: K Nearest Neighbour; RF: Random Trees; GBC: Gradient Boosting Classifier; ADA: AdaBoost; ETC: Extra Trees Classifier; SVC: Support Vector Classifier

#### Missing values

There was an average missingness of 3% (Supplementary figure 1). The most common missing variables were smoking status (16.1%), HbA1c (12.8%) and AST (10.3%). KNN Imputation was used as a pre-processing step to handle missing values in the whole dataset [14]. Other imputation methods considered were Mean/Median imputation, multiple imputation, Multiple Imputation by Chained Equations Imputation and Regression imputation. Our previous work demonstrated that KNN Imputation was superior in maintaining the integrity of the dataset, preserving both the distribution and correlation of variables whilst minimising the effect of missing variables on the performance [14].

#### ML model selection

The ID LIVER-ML development pipeline involved a comprehensive comparison of various machine learning models. More advanced techniques for hyperparameter tuning and deep learning modelling were used in the algorithm development compared to our previous work [12]. We compared both traditional ML algorithms and advanced ensemble methods ensuring a thorough evaluation of their performance on the dataset. Additionally regularised regression such as LASSO (Logistic Regression L1), Ridge (Logistic Regression L2) and Elastic Net were used to prove a nuanced understanding of logistic regression techniques.

#### Hyperparameter tuning

For hyperparameter tuning, we employed both grid search and random search methods to optimise the performance of each algorithm. Grid search systematically explores a specified subset of hyperparameters whilst random search samples a wide range of hyperparameters randomly with the benefits of being more efficient and effective. Details on the hyperparameters for each algorithm tuning methods and the range of values considered are in Supplementary Table 1.

#### Inclusion of Deep Learning Models

To extend our comparison, a simple feedforward neural network was used as a benchmark. Despite the potential for overfitting due to the size of the cohort and signal strength, this algorithm provided a point of reference for evaluating the performance of traditional ML models against a basic deep learning approach. The architecture and configuration of the feedforward neural network are detailed in Supplementary Table 2.

#### ML model evaluation

K-fold stratified cross-validation was used to compare the performance of various classification models (Supplementary Table 1) in cohort 1 (Figure 1). The CatBoost Classifier achieved the highest mean AUC score of 0.74 (95% CI 0.65-0.82). Logistic Regression (LR) and Linear Discriminant Analysis (LDA) also performed well with mean AUC scores of 0.73. Results for other models demonstrated competitive performance (Supplementary table 3). The CatBoost Classifier is an ML algorithm that uses gradient boosting and ordered boosting on decision trees to make predictions. The CatBoost Classifier has been shown to be effective in machine learning derived diagnostics [15].

#### Variable importance

The variable importance analysis method employed used a combination of different classifiers, including Random Forest, Extra Trees, AdaBoost, Gradient Boosting, and Support Vector Classifier. This analysis identified the most influential variables in their relative importance. Variable importance values specific to each classifier was obtained and aggregated to create a mean feature importance.

#### Determining the optimal cut off for model performance

Two methods were used to determine the optimal cut off threshold for ID LIVER-ML. Youden’s Index was used to identify the optimal cut off based off the ROC curve (Supplementary Figure 2). The optimal cut off using this method was 0.47. Secondly, F1 scores for each threshold were calculated from the precision-recall curve (Supplementary Figure 3). Using this method, the optimal cut off was 0.36. As the F1 score does not as effectively account for sensitivity and specificity that are critical in medical decision making, the cut off using Youden’s Index was used.

#### Calculation of conventional risk assessment scores

Validated fibrosis tests including FIB-4, NAFLD Fibrosis Score (NFS), AST to ALT ratio, AST to Platelet Ratio Index (APRI) and BARD score were calculated as per the published formulas for all patients [16–18] and compared to ID LIVER-ML using standard diagnostic tests at accepted age controlled thresholds [7, 8].

#### Subgroup analysis

The differing natural history of disease between aetiologies, age and, specifically, the presence of alcohol related fibrosis could affect the reliability of serum-based scores [19–21]. To ensure ID LIVER-ML can be applied to all patients, a subgroup analysis was undertaken on the following groups: 1) Patients with risk factors for alcohol-related liver disease (ARLD) compared to patients with purely metabolic risk factors; 2) the impact of number of risk factors; 3) age; 4) comparing the different TE thresholds. The Baveno VII guidelines use a cut off of 10kPa to rule out significant fibrosis, therefore we compared ID LIVER-ML performance using cut offs of either 8.0kPa or 10.0kPa [7].

## Results

### Cohort 1 analysis

Cohort 1 included 2039 patients with risk factors for liver disease. The cohort demographics are shown in Table 1. 1195 (58.2%) patients were male, 858 (41.8%) patients were female. 885 (43.1%) patients had type 2 diabetes and there was a mean BMI of 30.5 (SD ±6.2) kg/m^2^. The median alcohol consumption across the whole cohort was 24 grams per week. 515 (25.1%) patients had an abnormal ALT (>35IU/L in females, >50IU/L in males). 431 (21.1%) patients had a LSM greater than 8.0kPa. Cohort 1 was split randomly to create training, testing and validation 1 cohorts (Figure 1). There were no significant differences in the cohort demographics and laboratory results between the groups (Table 1).

**Table 1:** Characteristics of the Cohort 1: Training cohort, Testing cohort and hold out Validation cohort. Parametric data is presented as mean (standard deviation) with a three-way ANOVA used to assess significance between training, testing and validation 1 cohorts. Non-parametric data is presented as n (%) with the Kruskal-Wallis test used to assess significance between the three cohorts.

| Dataset characteristic | Cohort 1<br>(n=2039) | Training<br>(n= 1304) | Testing<br>(n=408) | Validation 1<br>(n=327) | P value:<br>training vs<br>testing vs<br>Validation 1 |
| --- | --- | --- | --- | --- | --- |
| **Age** (years), mean (SD) | 57.5 (14.2) | 57.6 (14.1) | 57.0 (14.2) | 57.6 (14.26) | 0.730 |
| **Gender**,<br>Male, n (%)<br>Female, n(%) | 1195 (58.2)<br>858 (41.8) | 765 (58.7)<br>539 (41.3) | 229 (56.1)<br>179 (43.9) | 187 (57.2)<br>140 (42.8) | 0.616 |
| **BMI** (kg/m2), mean (SD) | 30.5 (6.4) | 30.6 (6.5) | 30.6 (6.47) | 30.1 (5.9) | 0.377 |
| **Type 2 Diabetes**, n (%) | 882 (43.3) | 562 (43.1) | 167 (40.9) | 153 (46.8) | 0.276 |
| **Ethnicity**, n (%)<br>White<br>Black<br>South Asian<br>East Asian<br>Other | 1568 (76.4)<br>109 (5.3)<br>302 (14.7)<br>54 (2.6)<br>5 (0.2) | 1004 (77.0)<br>73 (5.6)<br>186 (14.3)<br>36 (2.8)<br>4 (0.3) | 314 (77.0)<br>24 (5.9)<br>58 (14.2)<br>11 (2.7)<br>1 (0.2) | 250 (76.5)<br>12 (3.7)<br>58 (17.7)<br>7 (2.1)<br>0 | 0.978 |
| **Alcohol** (g/week), median | 24 | 24 | 24 | 16 | 0.222 |
| **ALT** (IU/L), mean (SD) | 37.3 (29.7) | 37.2 (28.8) | 38.5 (34.3) | 35.9 (27.5) | 0.508 |
| **AST** (IU/L), mean (SD) | 35.0 (30.8) | 35.0 (29.2) | 37.2 (39.0) | 32.5 (24.9) | 0.150 |
| **ALP** (IU/L), mean (SD) | 88.5 (38.8) | 88.1 (39.2) | 89.3 (38.5) | 89.6 | 0.776 |
| **Albumin** (g/dL), mean (SD) | 40.1 (4.3) | 40.1 (4.3) | 39.9 (4.5) | 40.1 (3.9) | 0.591 |
| **Haemoglobin** (g/L) (SD) | 140.7 (15.1) | 140.4 (15.4) | 140.3 (14.2) | 142.1 (14.8) | 0.169 |
| **MCV** (fL), mean (SD) | 90.0 (10.0) | 90.0 (10.2) | 90.1 (11.6) | 89.8 (6.3) | 0.941 |
| **Platelet count** (x10<sup>9</sup>/L), mean (SD) | 249.7 (68.9) | 248.8 (66.9) | 249.5 (72.0) | 253.2 (73.1) | 0.591 |
| **Triglyceride** (mmol/L), mean (SD) | 1.8 (1.2) | 1.8 (1.2) | 1.8 (1.4) | 1.8 (1.2) | 0.623 |
| **HDL** (mmol/L), mean | 1.4 (0.5) | 1.4 (0.5) | 1.4 (0.5) | 1.4 (0.4) | 0.757 |
| (SD) |  |  |  |  |  |
| **Transient Elastography** (kPa), median | 7.3 (6.9) | 7.3 (6.9) | 7.3 (6.4) | 7.5 (7.9) | 0.897 |
| **IQR/Median** (%), mean | 13.2 (10.4) | 13.2 (9.5) | 13.2 (9.3) | 13.7 (14.5) | 0.769 |
| **At risk of fibrosis** (kPa>8.0), n (%) | 431 (21.1) | 270 (20.7) | 89 (21.8) | 68 (20.8) | 0.889 |

### Performance of ID LIVER-ML

Validation cohort 1 consisted of 327 patients. Consistent with the machine model derivation data, ID LIVER-ML demonstrated excellent performance in predicting the presence of clinically significant liver disease based on LSM (Training cohort: AUC 0.75 (95% CI 0.70-0.79), Validation 1 cohort: AUC 0.83 (95% CI 0.78 – 0.88)(Table 2). When predicting patients with a lower risk of fibrosis (LSM <8.0kPa), the model had a precision of 0.81 and Recall 0.94 (F1 score 0.87). When predicting patients with a risk of clinically significant liver disease (LSM >8.0kPa), precision and recall were not as accurate (0.64 and 0.33 respectively (F1 score 0.44)). Supplementary table 4 shows the prediction performance of ID LIVER-ML.

**Table 2:** Performance of ML algorithm and conventional scoring systems in the validation 1 dataset. Cut offs were chosen from current guidelines. APRI: AST to Platelet Ratio Index; NFS: NAFLD Fibrosis Score. DeLong test used to compare the performance of ID LIVER-ML to the conventional scoring systems.

| Model | Cut off | Sensitivity | Specificity | PPV | NPV | AUC (95% CI) | P value vs ML algorithm |
| --- | --- | --- | --- | --- | --- | --- | --- |
| **ML algorithm** | 0.47 | 0.71 | 0.78 | 0.46 | 0.91 | 0.83 (0.78-0.88) | - |
| **FIB-4 Score** | >1.3 | 0.57 | 0.65 | 0.30 | 0.85 | 0.65 (0.57-0.73) | **<0.001** |
| **BARD Score** | >2 | 0.82 | 0.32 | 0.24 | 0.87 | 0.61 (0.54-0.69) | **<0.001** |
| **APRI** | >0.7 | 0.23 | 0.96 | 0.58 | 0.82 | 0.65 (0.57-0.73) | **<0.001** |
| **NFS** | >-1.45 | 0.79 | 0.46 | 0.30 | 0.90 | 0.71 (0.65-0.78) | **<0.001** |
| **AST/ALT Ratio** | >1 | 0.33 | 0.65 | 0.18 | 0.78 | 0.53 (0.44-0.61) | **<0.001** |

### Description of variables

A ranking-based selection approach was employed using mean feature importance values to identify the key variables from 67 common variables in the SLP and ID LIVER datasets. Variables with higher mean importance values were prioritised for inclusion in the predictive model. Inclusion of these variables improved model performance metrics without significantly increasing the feature set. The top eight variables, as determined by their mean feature importance, which were used in ML algorithm development are AST (0.100), BMI (0.090), HbA1c (0.067), Platelet count (0.058), Triglycerides (0.052), ALT (0.052), HDL (0.042) and ALP (0.038) (Figure 3). Mean feature importance for all variables is available in Supplementary figure 4. Higher BMI, AST and HbA1c results were associated with increased predicted probabilities although a significant individual variability remained (Figure 4)

**Figure 3:**
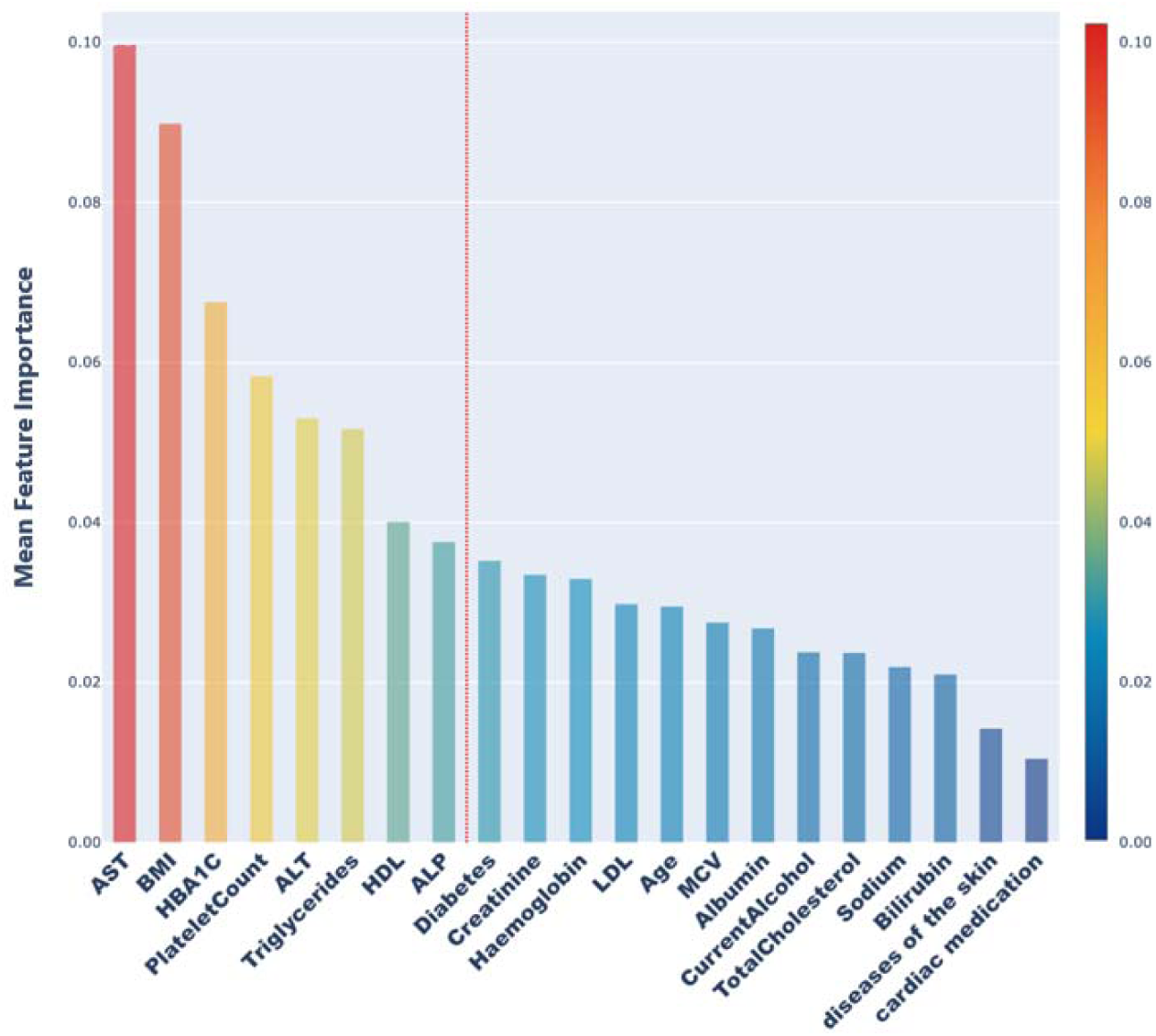
Variables with highest ML importance scores. The figure shows all clinical variables given a mean feature importance score of greater than 0.01 (21 of 67 variables analysed). Variables left of the red line were selected for use in ID LIVER-ML including: AST (0.100), BMI (0.090), HbA1c (0.067), Platelet count (0.058), Triglycerides (0.052), ALT (0.052), HDL (0.042) and ALP (0.038). Mean feature importance scores for all variables are available in Supplementary Figure 4.

**Figure 4:**
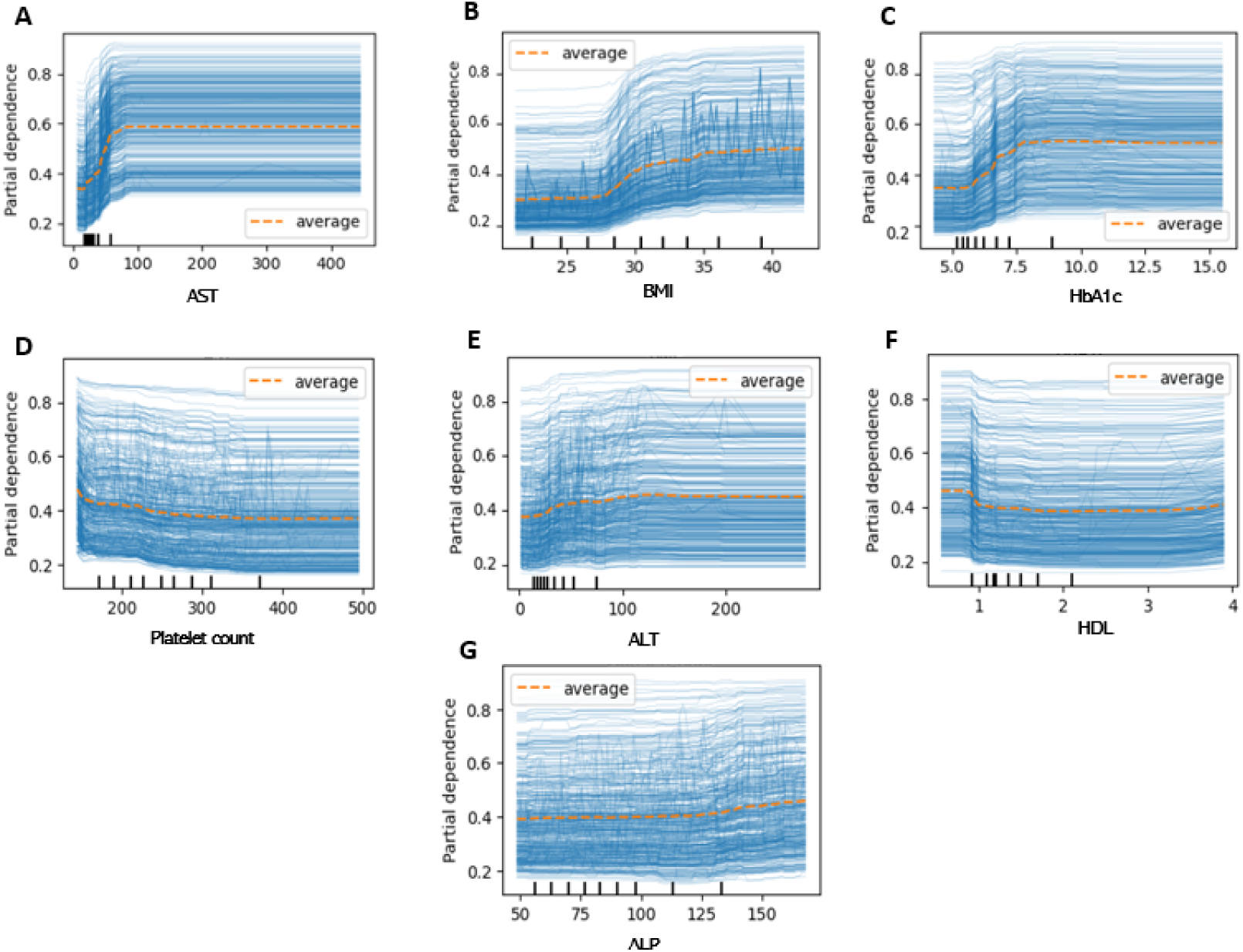
Relationship of each variable on the outcome. Partial Dependence Plots and Individual Conditional Expectation Plots for AST (A), BMI (B), HbA1c (C), Platelet count (D), ALT (E), HDL (F) and ALP (G). Higher results for AST, BMI and HbA1c were associated with increased predicted probabilities indicating their positive correlation with the outcome.

### Head-to-head comparison of ID LIVER-ML to conventional serum based scoring systems in Cohort 1

ID LIVER-ML performed well at predicting significant liver disease when applied to Validation 1 cohort (AUC 0.83 (95% CI 0.78-0.88))(Table 2). The commonly used risk stratification scores did not perform well on the real-world mixed aetiology cohorts (AUC_FIB-4_: 0.65 (95% CI 0.57-0.73); AUC_NFS_: 0.71 (95% CI 0.65-0.78); AUC_APRI_: 0.65 (95% CI 0.57-0.73); AUC_BARD_: 0.61 (95% CI 0.54-0.69); AUC_AST:ALT_: 0.53 (95% CI 0.44-0.61)) and were significantly outperformed by ID LIVER-ML (p_ML vs FIB-4_ <0.001; p_ML vs NFS_ <0.001; p_ML vs APRI_ <0.001; p_ML vs BARD_ <0.001, p_ML vs. AST:ALT_ <0.001)(Table 2; Supplementary Figure 5). In large scale screening, ruling out significant disease is an important economical consideration. ID LIVER-ML has a NPV (0.91) better than that of the FIB-4 score (0.85), APRI (0.82), BARD (0.87) and AST:ALT ratio (0.78). Only the NFS had a comparable NPV (0.90). Furthermore ID LIVER-ML had a comparable PPV to the traditional risk stratification scores. (Table 2).

### Subgroup analysis in Validation 1 cohort

Subgroup analysis of ID LIVER-ML was undertaken within the Validation 1 cohort in order to ensure equitable performance across key patient populations (Supplementary table 5).

### ML Algorithm performance by subgroup of risk factor for steatotic liver disease

Conventional risk stratification scores such as FIB-4 do not perform well in all patient groups, for example FIB-4 does not perform well in ARLD therefore ID LIVER-ML performance was assessed in all risk factor subgroups [22]. The algorithm performance remained good in patient subgroups with 1 risk factor (AUC_obesity_ 0.86 (95% CI 0.74 – 0.98), AUC_T2DM_ 0.82 (95% CI 0.70-0.94), AUC_alcohol_ 0.90 (0.80-1.00)) and performed better than FIB-4 score in metabolic risk factors (AUC_obesity_ 0.69 (95% CI 0.42-0.96)(p = 0.11), AUC_T2DM_ 0.50 (95% CI 0.30-0.70) (p = 0.001). There was no difference in the performance of the ID LIVER-ML and FIB-4 score in patients with an alcohol risk factor (FIB-4 AUC_alcohol_ 0.89 (95% CI 0.80-0.98) (p = 0.87)). Although the algorithm performance in patients with 2 metabolic risk factors was not as good, this remained better than the performance of the FIB-4 score (AUC_ID-LIVER ML_ 0.68 (95% CI 0.57-0.80) vs AUC_FIB-4_ 0.60 (95% CI 0.46-0.73) (p = 0.361)). In patients with a combination of metabolic and alcohol related risk factors although the performance of ID LIVER-ML was reduced, it continued to out-perform the FIB-4 score however did not reach statistical significance (AUC_MetALD_: 0.74 (95% CI 0.52 – 0.96) vs 0.66 (95% CI 0.41 – 0.91) (p = 0.614)).In patients without T2DM, the ID-LIVER ML maintained good performance (AUC_noT2DM_ 0.86 (95% CI 0.77-0.94)).

### ML Algorithm performance by age

ID LIVER-ML performed better in patients under 65 (AUC_Age <65_ 0.87 (95% CI 0.82 – 0.92) than those over 65 (AUC _Age >=65_ 0.79 (95% CI 0.70-0.89) (p=0.002). Given the well documented differences in FIB-4 cut off in age [21], a FIB-4 cut off of 2.00 was used in patients over 65. FIB-4 performed significantly better in patients <65 than those >65 (AUC_AGE<65_ 0.70 (95% CI 0.60 – 0.80) vs AUC_AGE>=65_ 0.55 (95% CI 0.40-0.70))(p = 0.03). ID LIVER-ML outperformed the FIB-4 score in both age categories (AUC_<65_ 0.87 vs 0.70 (p = 0.002); AUC_>=65_ 0.79 vs 0.55 (p = 0.016)).

### ML algorithm performance at predicting LSM >10.0kPa

Although ID LIVER-ML was trained to predict the risk of clinically significant liver disease using a LSM cut off of 8.0kPa, it remained stable when a higher LSM cut off (10kPa) was applied (Sensitivity 0.67, Specificity 0.74, AUC 0.80 (95% CI 0.74 – 0.87)). ID LIVER-ML significantly out-performed the FIB-4 score (AUC_ML_ 0.80 vs AUC_FIB-4_ 0.66)(p = 0.007).

### Comparison of FIB4 and ID LIVER-ML as a community screening tool Methodology

The prospective validation cohort, “Validation 2” consisted of 399 patients. 19 patients were excluded as a FIB-4 score could not be calculated due to missing variables resulting in a total cohort of 380 patients with metabolic, alcohol related or mixed aetiologies (Figure 1).

### Cohort Characteristics

The Validation 2 dataset consisted of a different population to the validation 1 cohort. Cohort characteristics are presented in Table 3. The cohort consisted of 203 (53.4%) males. The mean age was 56.4 (±12.2) years. 145 (38.2%) patients had a diagnosis of T2DM and the mean BMI was 32.8 (±6.2) kg/m^2^. 123 (32.4%) patients had a raised ALT. 53 (14.0%) patients drank harmful quantities of alcohol and 201 (52.9%) patients declared no alcohol use. 46 (12.0%) patients had liver stiffness measurements greater than 8.0 kPa.

**Table 3:**
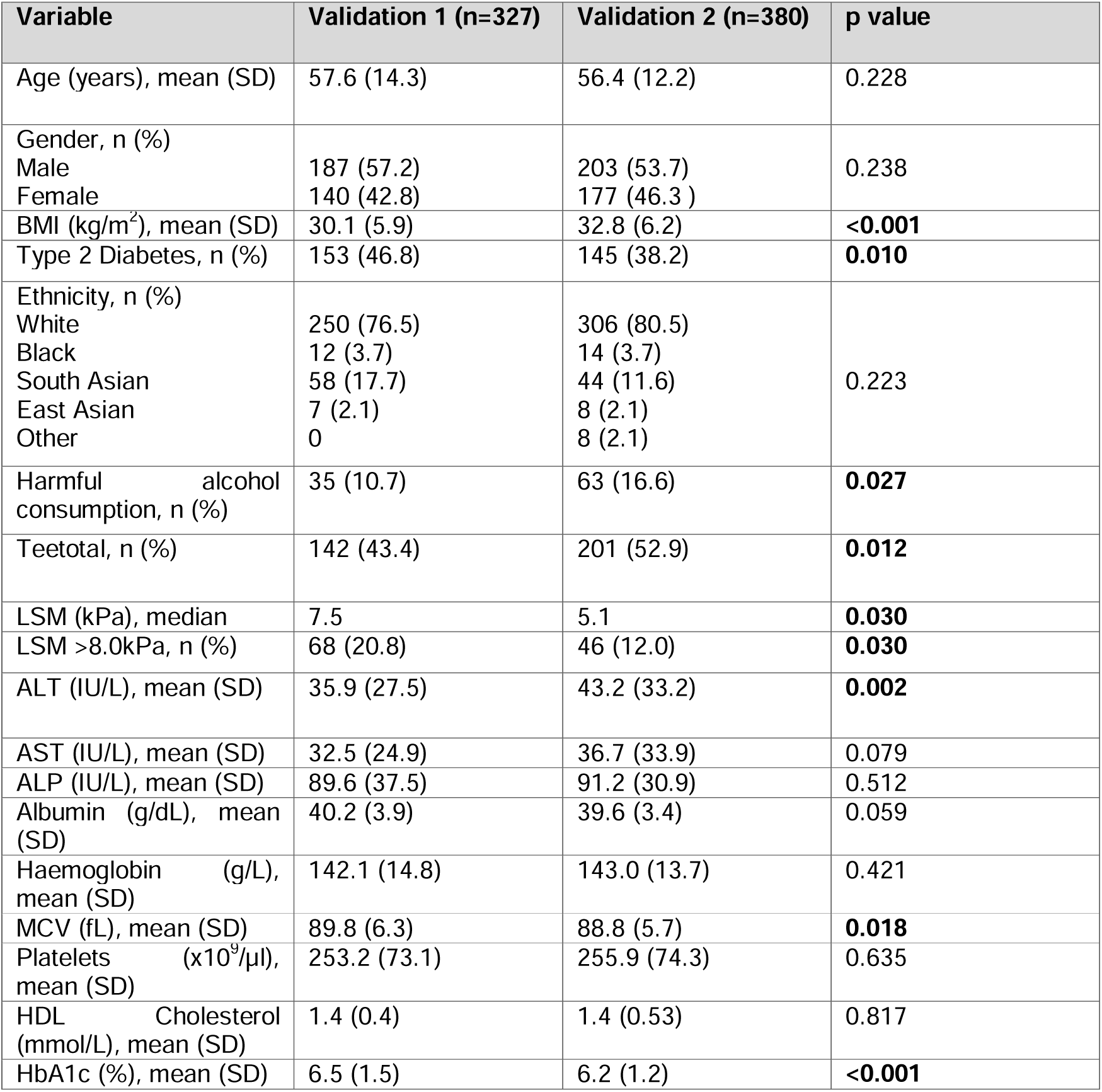
Clinical demographics of Validation 2 cohort compared to the validation 1 cohort. Parametric data is presented as mean (standard deviation) with Student’s T Test used to assess significance between validation 1 and validation 2 cohorts. Non-parametric data is presented as n (%) with the Mann Whitney U test used to assess significance between the two cohorts.

### Performance of the FIB-4 score and ML algorithm and outcomes within a clinical pathway

In the Validation 2 dataset, ID LIVER-ML remained stable and outperformed FIB-4 score at detecting patients at risk of having clinically significant liver fibrosis (AUC_ID LIVER-ML_ 0.85 (95% CI 0.79-0.91) vs AUC_FIB-4_ 0.78 (0.70-0.86)) (Supplementary Figure 6).

FIB-4 score performed significantly better in the Validation 2 cohort than it did in the Validation 1 cohort (p=0.02). In comparison to the patients in Validation 1 cohort, patients in Validation 2 cohort had a significantly larger mean BMI (32.8kg/m^2^ vs 30.1kg/m^2^ (p = <0.001; 95% CI 1.81 – 3.60)). The proportion of patients with T2DM was significantly larger in the Validation 1 cohort (46.8% vs 38.2% (p = 0.01)) with a higher mean HbA1c (6.5% vs 6.2% (p = <0.001; 95% CI -0.61 - -0.18). Validation 2 cohort contained a higher proportion of patients who drank harmful quantities of alcohol (16.6% vs 10.7% (p = 0.027)) or who had no declared alcohol consumption (52.9% vs 43.4% (p = 0.012)) compared to Validation 1 cohort.

Extrapolating this to a possible clinical pathway (Supplementary Figure 7) using the FIB-4 score and current clinical guidelines, 142/380 (37.4%) patients required further fibrosis assessment through TE. Of these 34 patients (23.9%) had clinically significant liver disease on TE requiring further investigations in Hepatology clinic. Using ID LIVER-ML, significantly fewer patients (60 (15.8%) would require further fibrosis assessment (Figure 5) with a subsequent 30 (50%) patients found to have clinically significant liver disease on TE (p=<0.001). Therefore significantly fewer patients had unnecessary investigations if ID LIVER-ML was used compared to if the FIB-4 score was used (p = <0.001). There were no significant differences in false negative outcomes between the FIB-4 score (12/239 (5.0%)) and ID LIVER-ML (17/320 (5.3%))(p=0.712).

**Figure 5:**
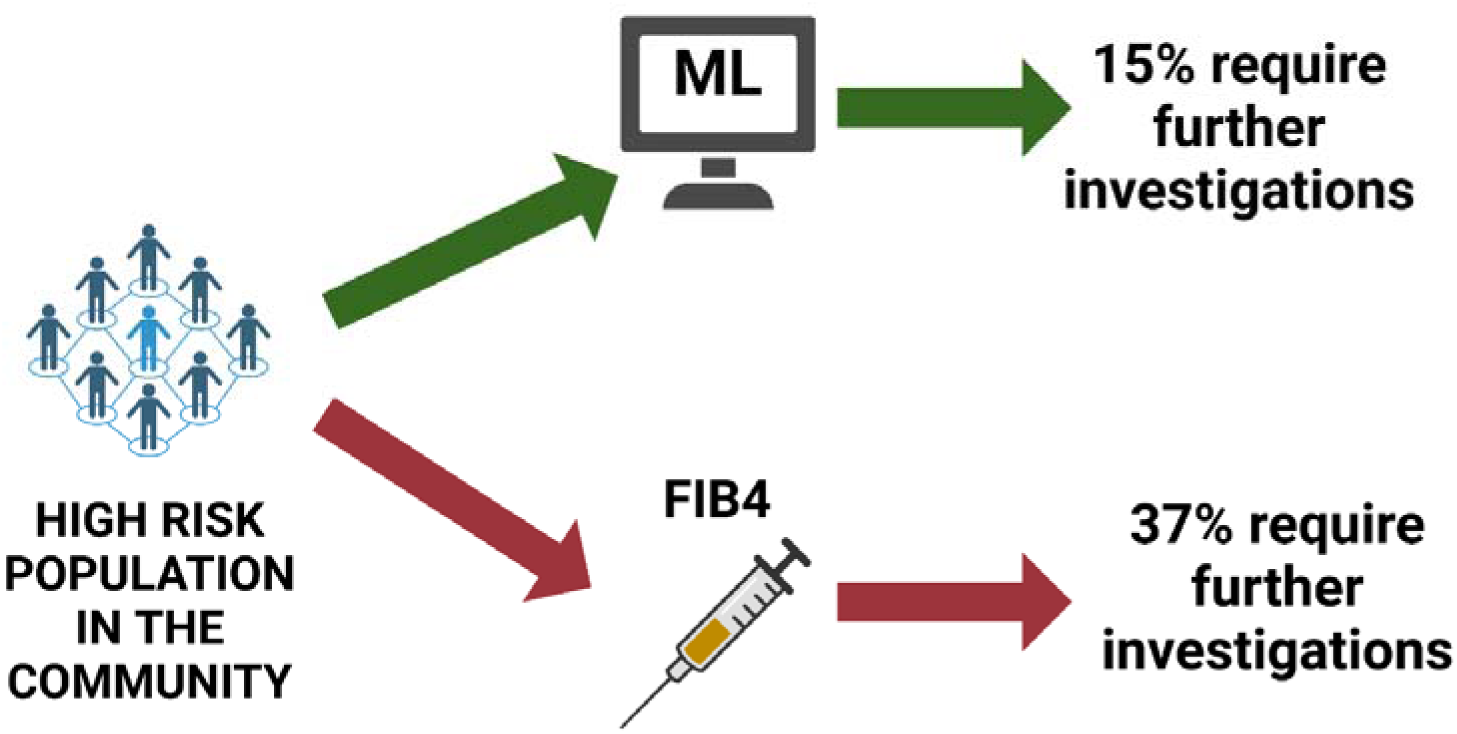
Diagram demonstrating the differences in number of patients requiring further investigations when ID LIVER-ML is used compared to the FIB-4 score.

Expanding this over a larger population, we identified 55,286 patients with at least one risk factor for liver disease using SNOMED codes from a screened population of 432,201 patients in Primary Care Practices in Manchester, UK. Therefore, extrapolating the clinical outcomes across the at-risk population, using ID LIVER-ML would save 12,384 patients from unnecessary investigations. Economic analysis has been performed separately and shows that the ID LIVER-ML algorithm would meet the English cost-effectiveness thresholds (£20,000/QALY gained) with a reasonable cost of £10,068/QALY gained [23].

## Discussion

Steatotic liver disease is highly prevalent in the population however there are concerns that traditional risk stratifications scores can underperform in population based screening. Our study shows that ID LIVER-ML predicts the risk of clinically significant liver fibrosis in a mixed cohort of patients with risk factors for steatotic liver disease in a primary care setting with a significantly better performance than traditional blood based risk scoring systems.

The use of risk stratification scores such as FIB-4 and NFS is widespread in clinical practice and is recommended in UK and European guidelines [7, 8, 24]. They have notable benefits in that they are inexpensive and use routine clinical information that is easy to apply in primary care. Furthermore they have a persistently good NPV as demonstrated in our study, and can accurately stratify the risk of cirrhosis decompensation or hepatocellular carcinoma [25]. They do, however, have limitations. FIB-4 score has been shown to be less effective in patients over 65 years old, patients with alcohol related risk factors and in patients with T2DM [21, 22, 25]. Despite their good NPV, FIB-4 and NFS have a poor PPV resulting in considerable clinical and economic impact in constrained healthcare systems.

ID LIVER-ML is designed to be utilised as a primary care tool as a first gate, superseding the traditional risk stratification score. It can be automated within large primary care datasets and electronic health records to distinguish at-risk individuals. ID LIVER-ML has similar advantages to FIB-4 and NFS, with fewer disadvantages. Its superior specificity to the FIB-4 score and NPV comparable with NFS makes ID LIVER-ML a better tool to predict clinically significant liver disease. It performed well across all age groups and different aetiologies making it more equitable than the traditional risk stratification scores. Although additional blood tests are required for ID LIVER-ML, the current pathway requires this to calculate a FIB-4 score, and tests such as lipid profile and HbA1c should be performed as part of a non-invasive liver aetiological screen and subsequent cardiovascular risk assessment anyway. These have the potential to be reflex tested [26].

ID LIVER-ML was built using a real-world dataset consisting of patients with a mixed range of risk factors for liver disease. FIB-4 and NFS were designed using biopsy proven secondary care cohorts [16, 17]. The changes in nomenclature emphasises a spectrum in steatotic liver disease from MASLD through MetALD to ARLD [27]. There is a high prevalence of patients with mixed aetiology and high disease burden resulting in a high pre-test probability. Therefore, there is the need for tests that accurately stratify risk that work in cohorts with a mixed aetiology rather than just a single aetiology.

ML has been used before to predict the risk of clinically significant liver disease. The LiverAID models confirm the ability of ML to accurately predict the risk of liver disease within a primary care setting [28]. However, some variables would not be routinely collected or available in primary care such as mid-upper arm circumference which is required for all but the least complex models. Unlike other models, ID LIVER-ML is trained on a solely primary care recruited population with risk factors undergoing initial screening. Its performance is similar to models developed on biopsy proven cohorts, such as in the development of the SAFE screening tool, or in the general population such as the LiverRisk score [29, 30].

Other novel solutions such as iLFTs involve reflex testing to risk stratify patients in primary care who may need secondary care input [27]. Although these use existing risk stratification scores, there could be potential for machine learning solutions such as ID LIVER-ML to be incorporated within a reflex testing solution within primary care.

The main strength of ID LIVER-ML lies in training on real-world cohorts in two UK cities with a heterogeneity of aetiologies. As a result, ID LIVER-ML is highly applicable for patients with varying numbers of risk factors as well as risk factors across the spectrum of steatotic liver disease. Patients with mixed aetiologies are often missed from non-real-world cohorts yet are an important and large group (29.5% of Validation 2 cohort had MetALD). Using real-world data from two different UK cities reduces the risk that the model will not be universally applicable. We used LSM as a surrogate marker of clinically significant liver fibrosis and as the ground truth for ID LIVER-ML. This increases the applicability to a primary care population. LSM is widely used for fibrosis assessment over biopsy and screening high-risk patients in a pre-hospital setting using biopsy would not be feasible.

Limitations of the study include the LSM threshold used to determine clinically significant liver disease. The manufacturer’s cut-offs for MASLD and ARLD differ, with ARLD having a higher cut off for F2/F3 fibrosis than that of MASLD. Therefore, patients with alcohol related aetiologies may be determined to be at high risk of fibrosis and recommended for unnecessary further investigations. It should be noted when ID LIVER-ML was assessed against a higher cut-off of 10.0kPa, it maintained stability and outperformed FIB-4.

A potential limitation of ID LIVER-ML is the requirement of fasting lipids and HbA1c. This potentially predisposes bias towards patients with MASLD. This reflects the high prevalence of patients with MetALD in our real-world population and ID LIVER-ML performs well in patients with alcohol as a risk factor. This is in keeping with previously published models where triglycerides and HbA1c have been used to predict the risk of fibrosis in steatotic liver disease [28].

Finally, Machine learning is biased towards its training dataset. Most recruited patients were diagnosed with MASLD. Although our dataset is real-world data, the small proportion of patients with ARLD may not be representative of the general population. All of our recruited patients were deemed to be “at risk” as would be the norm in the current diagnostic pathway. Mitigating this would require a full screening programme.

To be an implementable solution for healthcare, further validation in external cohorts to demonstrate diagnostic performance stability.

## Conclusion

Machine Learning tools have great potential to improve quality and efficiency in population based testing for disease. We developed and deployed ID LIVER-ML to successfully predict the presence of clinically significant liver disease in a real-world cohort of patients with risk factors for steatotic liver disease of all aetiologies. Using readily available parameters, ID LIVER-ML out-performed conventional testing strategies and could be a cost-effective tool to stratify patients with risk factors for liver disease.

## Author Contributions

HP and LB jointly wrote the manuscript with contributions from MM and are joint first authors. HP curated the ID LIVER dataset. LB curated the SLP dataset. MM performed the ML work. RH, JM, ING and VSA advised on the creation of ID LIVER-ML. Internal review was done by SL, CM, OS, RH, MP, JM, JS, KPH, NH, ING and VSA. ING and VSA are joint senior authors.

## Ethics

Clinical Trials number: NCT 04666402 IRAS: 273633

REC reference: 20/NS/0055

## Data Availability

All data produced in the present study are available upon reasonable request to the authors

## Acknowledgments

We would like to acknowledge the UK Government’s Innovate UK Industrial Strategy Challenge Fund for funding and supporting this project.

## ID-LIVER Consortium

NAH is Chief Investigator of ID-LIVER.

Member organisations of the consortium, in alphabetical order, are: The British Liver Trust, GE Healthcare, Health Innovation Manchester, Jiva.ai, Manchester University NHS Foundation Trust, NorthWest EHealth, Nottingham University Hospitals NHS Trust, Octopus Ventures, Perspectum Diagnostics, Roche Diagnostics, Sectra, Sollis, The University of Manchester, The University of Nottingham, TRUSTECH and Vocal.

