## Supplementary data for "Improving the detection of clinically significant steatotic liver disease using a machine learning algorithm in a real-world primary care population"

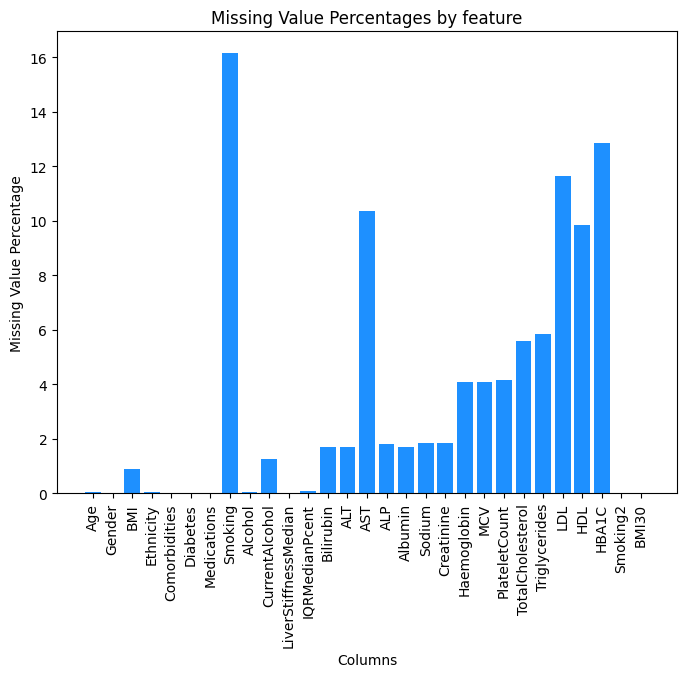

**Supplementary Figure 1: Bar chart showing percentage of each variable across cohort 1. The mean missingness across the whole cohort is <10%.**

**Supplementary Table 1: Hyperparameters for each model, tuning methods and the range and choices of values considered.**

| **Model** | **Hyperparameters** | **Tuning Method** | **Hyperparameter Range/Values** |
| --- | --- | --- | --- |
| **CatClassifier** | **depth, learning_rate** | **Random Search** | **depth: [3, 10], learning_rate: [0.01, 0.1]** |
| **Logistic Regression (LR)** | **C** | **Grid Search** | **C: [0.01, 0.1, 1, 10, 100]** |
| **Linear Discriminant Analysis (LDA)** | **solver** | **Grid Search** | **solver: [svd, lsqr, eigen]** |
| **XGBoost (XGB)** | **n_estimators, max_depth** | **Random Search** | **n_estimators: [100, 500], max_depth: [3, 10]** |
| **Gradient Boosting Classifier (GBC)** | **learning_rate, n_estimators** | **Grid Search** | **learning_rate: [0.01, 0.1], n_estimators: [100, 200]** |
| **Extra Trees Classifier (ETC)** | **n_estimators, max_depth** | **Random Search** | **n_estimators: [100, 500], max_depth: [None, 10]** |
| **Random Trees (RT)** | **n_estimators, max_features** | **Random Search** | **n_estimators: [100, 500], max_features: [auto, sqrt]** |
| **AdaBoost (Ada)** | **n_estimators, learning_rate** | **Grid Search** | **n_estimators: [50, 100], learning_rate: [0.01, 0.1]** |
| **Support Vector Machine (SVM)** | **C, kernel** | **Grid Search** | **C: [0.1, 1, 10], kernel: [linear, rbf]** |
| **Stacking Classifier (Ensemble)** | **base_estimators, final_estimator** | **Grid Search** | **base_estimators: [LR, SVM], final_estimator: [LR, GBC]** |
| **Logistic Regression (L1)** | **C** | **Grid Search** | **C: [0.01, 0.1, 1, 10, 100]** |
| **Logistic Regression (L2)** | **C** | **Grid Search** | **C: [0.01, 0.1, 1, 10, 100]** |
| **Logistic Regression (Elastic Net)** | **l1_ratio, C** | **Grid Search** | **l1_ratio: [0.1, 0.5, 0.9], C: [0.01, 0.1, 1, 10, 100]** |
| **FeedForward NN (Deep Learning)** | **layers, units per layer, activation** | **Random Search** | **layers: [2, 3], units per layer: [64, 128], activation: [relu, tanh]** |

**Supplementary Table 2: The architecture and configuration of the feedforward neural network**

| **Parameter** | **Value** |
| --- | --- |
| **Number of Layers** | **3** |
| **Units per Layer** | **128, 64, 32** |
| **Activation Function** | **ReLU** |
| **Optimizer** | **Adam** |
| **Loss Function** | **Binary Crossentropy** |
| **Epochs** | **50** |
| **Batch Size** | **32** |
| **Learning Rate** | **0.001** |
| **Regularization** | **Dropout (0.5)** |

**Supplementary Table 3: The performance of different machine learning models at predicting the presence of clinically significant liver disease (Cohort 1: Training dataset)**

| Classifier | ROC_AUC | 95% CI Lower | 95% CI Upper |
| --- | --- | --- | --- |
| CatBoost Classifier | 0.739437 | 0.652528 | 0.826347 |
| Logistic Regression (LR) | 0.736170 | 0.650129 | 0.822210 |
| Linear Discriminant Analysis (LDA) | 0.734859 | 0.646648 | 0.823070 |
| XGBoost (XGB) | 0.728634 | 0.648336 | 0.808932 |
| Gradient Boosting Classifier (GBC) | 0.730841 | 0.653780 | 0.807903 |
| Extra Trees Classifier (ETC) | 0.736825 | 0.645234 | 0.828417 |
| Random Trees (RT) | 0.736457 | 0.651035 | 0.821879 |
| AdaBoost (Ada) | 0.732122 | 0.644337 | 0.819908 |
| Support Vector Machine (SVM) | 0.736577 | 0.655124 | 0.818029 |
| Stacking Classifier (Ensemble) | 0.678172 | 0.623795 | 0.732549 |
| Logistic Regression (L1) | 0.629902 | 0.629902 | 0.629902 |
| Logistic Regression (L2) | 0.644608 | 0.644608 | 0.644608 |
| Logistic Regression (Elastic Net) | 0.644608 | 0.644608 | 0.644608 |
| FeedForward NN (Deep Learning) | 0.557334 | 0.557334 | 0.557334 |

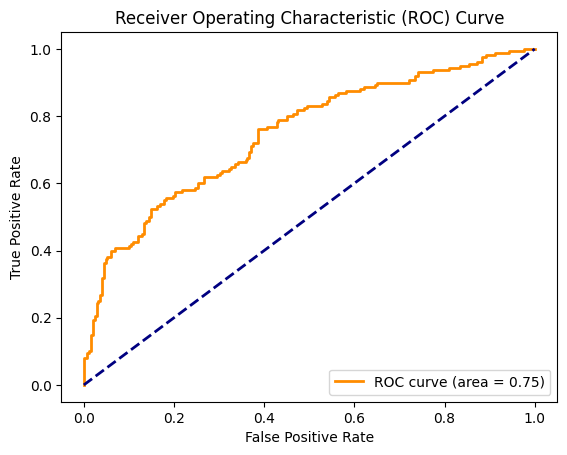

**Supplementary Figure 2: ROC Curve displaying the true positive rate (TPR) versus the false positive rate (FPR) across different thresholds.**

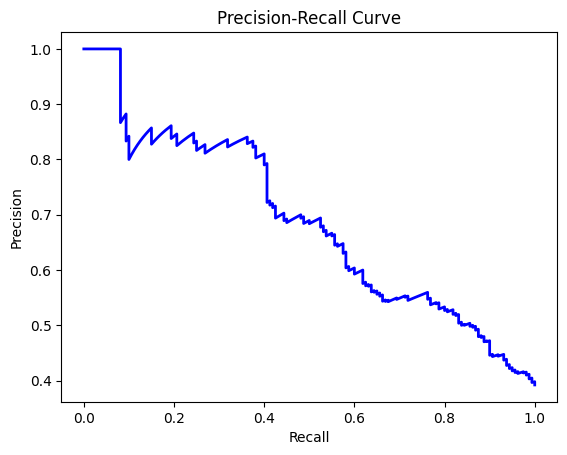

**Supplementary Figure 3: Precision-Recall Curve displaying precision versus recall across different thresholds.**

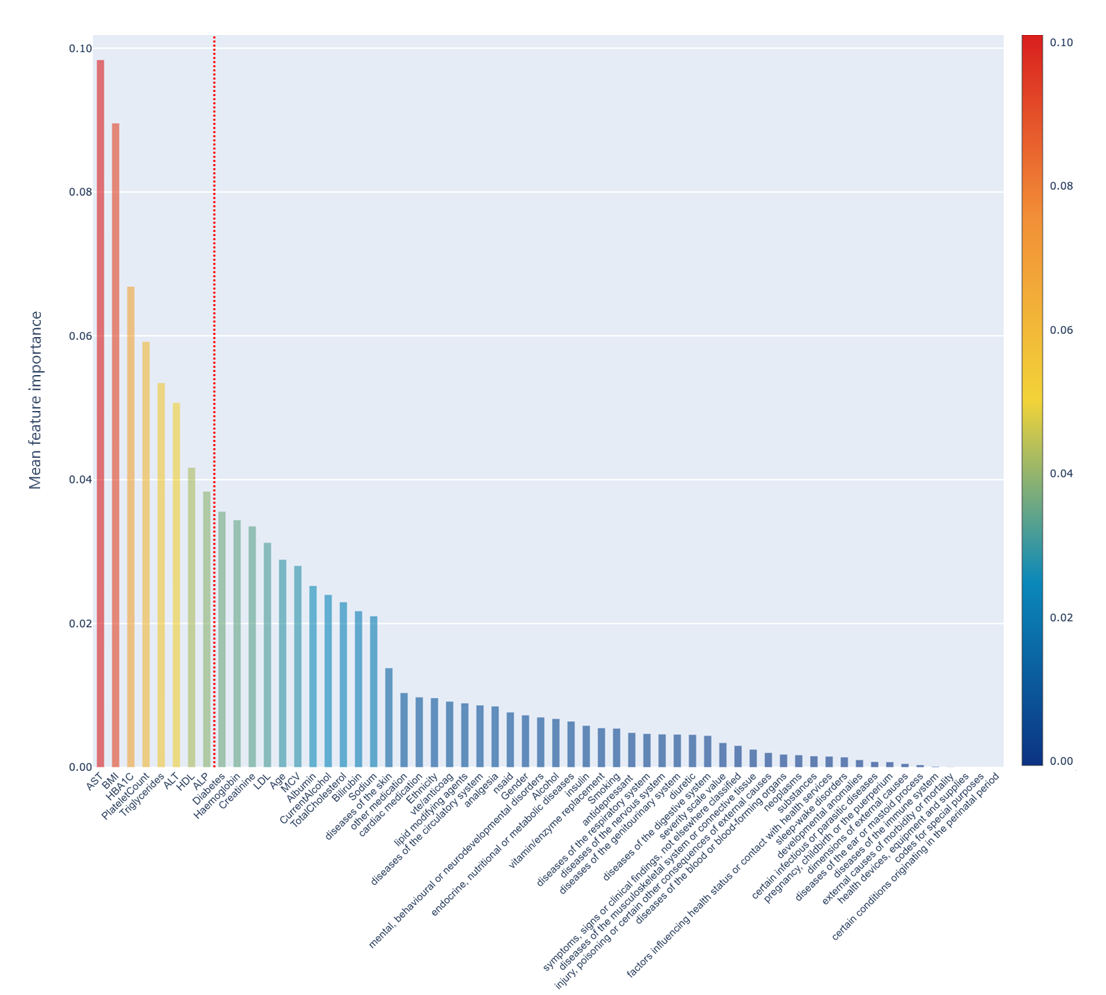

**Supplementary Figure 4: Mean feature importance for all common variables in the SLP and ID LIVER cohorts.** Variables to the left of the red line were used in the final ML model.

**Supplementary table 4: Performance of the CatBoost classifier in the validation 1 dataset.**

| Presence of Clinically Significant Liver Disease | LSM <8.0kPa | LSM >8.0kPa | Precision | Recall | F1 Score |
| --- | --- | --- | --- | --- | --- |
| Predicted Negative | 0.89 | 0.82 | 0.81 | 0.94 | 0.87 |
| Predicted Positive | 0.48 | 0.62 | 0.64 | 0.33 | 0.44 |

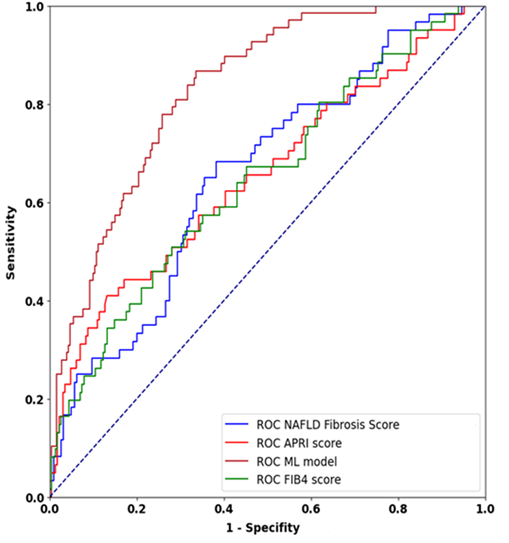

**Supplementary Figure 5: Receiver Operator Curve showing the performance of FIB-4 (green), APRI (red) and NFS (blue) compared to the ML model (Brown).** ML model AUC: 0.83, NAFLD Fibrosis Score AUC: 0.71, APRI AUC 0.65, FIB-4 AUC: 0.65.

**Supplementary table 5: Table showing the breakdown of performance of the CatBoost Classifier and the FIB-4 score by risk factor.** AUCs of the FIB-4 score and algorithm are compared using Delong’s Test.

| Subset | Method | Sensitivity | Specificity | PPV | NPV | AUC (95% CI) | P value |
| --- | --- | --- | --- | --- | --- | --- | --- |
| 3 RF = Alcohol + T2DM + obesity | FIB4 | 0.57 | 0.50 | 0.57 | 0.50 | 0.53 (0.30 - 0.77) | 0.56 |
|  | ML | 0.86 | 0.23 | 0.54 | 0.60 | 0.63 (0.41 - 0.85) |  |
| 1 RF = T2DM | FIB4 | 0.33 | 0.63 | 0.14 | 0.84 | 0.50 (0.30 – 0.70) | **0.001** |
|  | ML | 0.50 | 0.92 | 0.55 | 0.90 | 0.82 (0.70 - 0.94) |  |
| 1 RF = Obesity only | FIB4 | 0.40 | 0.85 | 0.33 | 0.88 | 0.69 (0.42 - 0.96) | 0.11 |
|  | ML | 0.67 | 0.78 | 0.52 | 0.86 | 0.86 (0.74 - 0.98) |  |
| 1 RF = Alcohol only | FIB4 | 1.00 | 0.64 | 0.23 | 1.00 | 0.89 (0.80-0.98) | 0.87 |
|  | ML | 0.75 | 0.94 | 0.60 | 0.97 | 0.90 (0.80-1.00) |  |
| 2 RF = Alcohol + Obesity | FIB4 | 0.55 | 0.58 | 0.38 | 0.73 | 0.60 (0.43 - 0.77) | **0.065** |
|  | ML | 0.76 | 0.60 | 0.47 | 0.84 | 0.77 (0.65 - 0.90) |  |
| 2 RF = T2DM and Obesity | FIB4 | 0.54 | 0.62 | 0.51 | 0.65 | 0.58 (0.47 - 0.70) | 0.63 |
|  | ML | 0.89 | 0.24 | 0.46 | 0.75 | 0.55 (0.35 - 0.72) |  |
| 2 RF = T2DM and Alcohol | FIB4 | 0.60 | 0.47 | 0.53 | 0.54 | 0.58 (0.36 - 0.79) | 0.47 |
|  | ML | 0.80 | 0.41 | 0.54 | 0.70 | 0.68 (0.50 - 0.88) |  |
| >=65 years old | FIB4 | 0.57 | 0.41 | 0.23 | 0.76 | 0.55 (0.40 - 0.70) | **0.016** |
|  | ML | 0.54 | 0.82 | 0.48 | 0.86 | 0.79 (0.70 - 0.89) |  |
| <65 years old | FIB4 | 0.55 | 0.75 | 0.36 | 0.87 | 0.70 (0.60 - 0.80) | **0.002** |
|  | ML | 0.79 | 0.76 | 0.45 | 0.94 | 0.87 (0.82 - 0.92) |  |
| LSM >10kPa | FIB4 | 0.56 | 0.63 | 0.20 | 0.90 | 0.66 (0.58 - 0.74) | **0.007** |
|  | ML | 0.67 | 0.74 | 0.30 | 0.93 | 0.80 (0.74 - 0.87) |  |

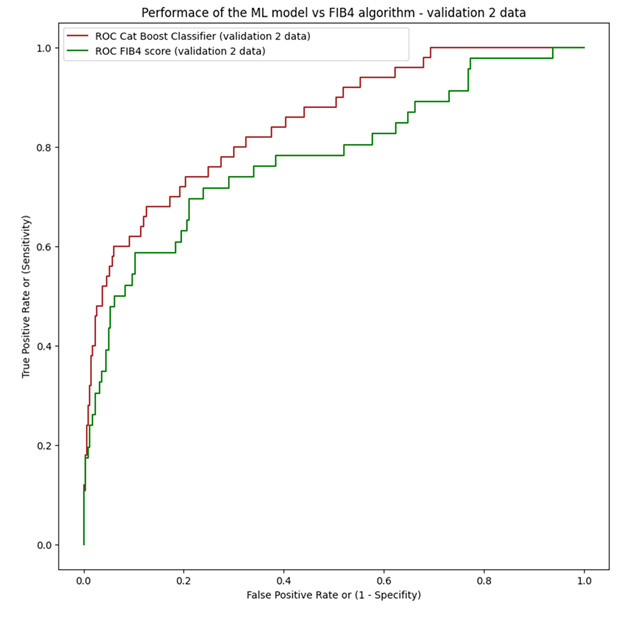

**Supplementary figure 6: performance of the ML classifier vs FIB4 in a validation group 2**

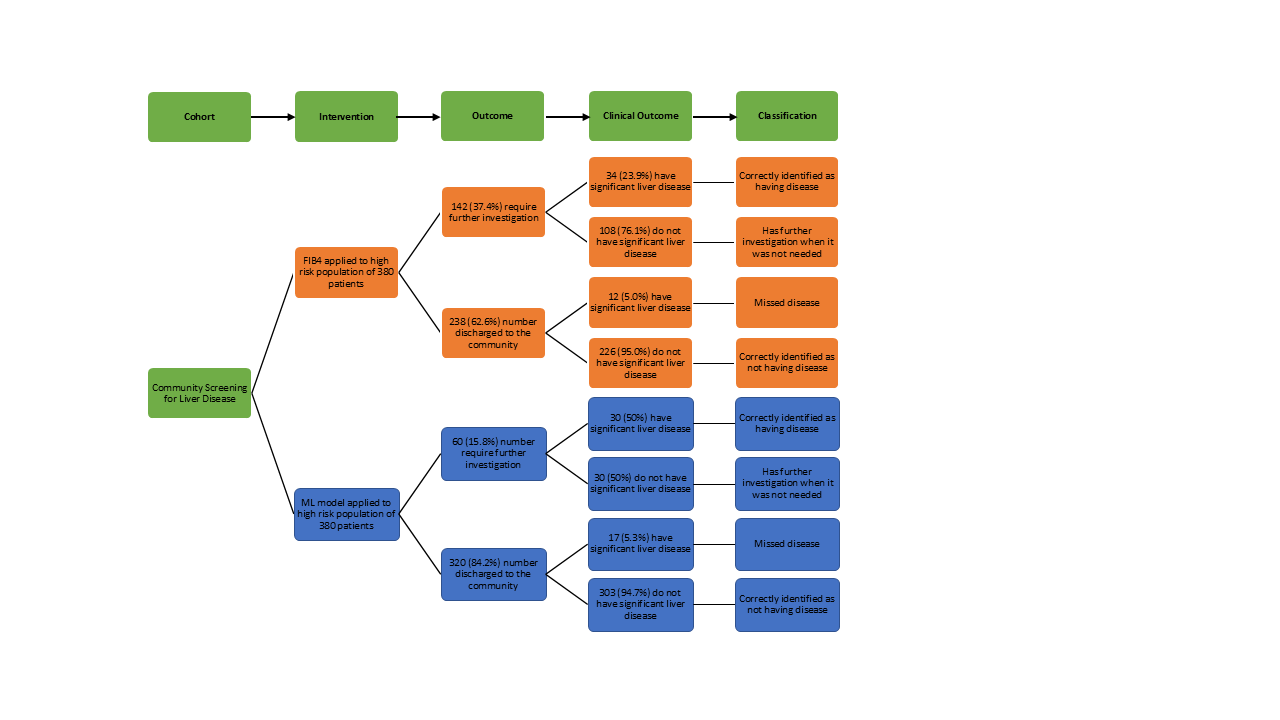

**Supplementary Figure 7 : Clinical outcomes from the application of the ML model to the Validation 2 population compared to FIB-4 score.**

*Transparent reporting – Tripod - https://www.tripod-statement.org/*

| **Section/Topic** | **Item** |  | **Checklist Item** | **Page** |
| --- | --- | --- | --- | --- |
| **Title and abstract** | | | | |
| Title | 1 | D;V | Identify the study as developing and/or validating a multivariable prediction model, the target population, and the outcome to be predicted. | 1 |
| Abstract | 2 | D;V | Provide a summary of objectives, study design, setting, participants, sample size, predictors, outcome, statistical analysis, results, and conclusions. | 2 |
| **Introduction** | | | | |
| Background and objectives | 3a | D;V | Explain the medical context (including whether diagnostic or prognostic) and rationale for developing or validating the multivariable prediction model, including references to existing models. | 4 |
|  | 3b | D;V | Specify the objectives, including whether the study describes the development or validation of the model or both. | 5 |
| **Methods** | | | | |
| Source of data | 4a | D;V | Describe the study design or source of data (e.g., randomized trial, cohort, or registry data), separately for the development and validation data sets, if applicable. | 5-6 |
|  | 4b | D;V | Specify the key study dates, including start of accrual; end of accrual; and, if applicable, end of follow-up. | 5-6 |
| Participants | 5a | D;V | Specify key elements of the study setting (e.g., primary care, secondary care, general population) including number and location of centres. | 5-6 |
|  | 5b | D;V | Describe eligibility criteria for participants. | 5-6 |
|  | 5c | D;V | Give details of treatments received, if relevant. | NA |
| Outcome | 6a | D;V | Clearly define the outcome that is predicted by the prediction model, including how and when assessed. | 5-6 |
|  | 6b | D;V | Report any actions to blind assessment of the outcome to be predicted. | NA |
| Predictors | 7a | D;V | Clearly define all predictors used in developing or validating the multivariable prediction model, including how and when they were measured. | 6-11 |
|  | 7b | D;V | Report any actions to blind assessment of predictors for the outcome and other predictors. | NA |
| Sample size | 8 | D;V | Explain how the study size was arrived at. | 5-6 |
| Missing data | 9 | D;V | Describe how missing data were handled (e.g., complete-case analysis, single imputation, multiple imputation) with details of any imputation method. | 10 |
| Statistical analysis methods | 10a | D | Describe how predictors were handled in the analyses. | 11-12 |
|  | 10b | D | Specify type of model, all model-building procedures (including any predictor selection), and method for internal validation. | 11-13 |
|  | 10c | V | For validation, describe how the predictions were calculated. | 13 |
|  | 10d | D;V | Specify all measures used to assess model performance and, if relevant, to compare multiple models. | 13 |
|  | 10e | V | Describe any model updating (e.g., recalibration) arising from the validation, if done. | NA |
| Risk groups | 11 | D;V | Provide details on how risk groups were created, if done. | NA |
| Development vs. validation | 12 | V | For validation, identify any differences from the development data in setting, eligibility criteria, outcome, and predictors. | 13 |
| **Results** | | | | |
| Participants | 13a | D;V | Describe the flow of participants through the study, including the number of participants with and without the outcome and, if applicable, a summary of the follow-up time. A diagram may be helpful. | 7 |
|  | 13b | D;V | Describe the characteristics of the participants (basic demographics, clinical features, available predictors), including the number of participants with missing data for predictors and outcome. | 13-14 |
|  | 13c | V | For validation, show a comparison with the development data of the distribution of important variables (demographics, predictors and outcome). |  |
| Model development | 14a | D | Specify the number of participants and outcome events in each analysis. | 8 |
|  | 14b | D | If done, report the unadjusted association between each candidate predictor and outcome. | NA |
| Model specification | 15a | D | Present the full prediction model to allow predictions for individuals (i.e., all regression coefficients, and model intercept or baseline survival at a given time point). | NA |
|  | 15b | D | Explain how to the use the prediction model. | NA |
| Model performance | 16 | D;V | Report performance measures (with CIs) for the prediction model. | 14 |
| Model-updating | 17 | V | If done, report the results from any model updating (i.e., model specification, model performance). | NA |
| **Discussion** | | | | |
| Limitations | 18 | D;V | Discuss any limitations of the study (such as nonrepresentative sample, few events per predictor, missing data). | 23 |
| Interpretation | 19a | V | For validation, discuss the results with reference to performance in the development data, and any other validation data. | 22 |
|  | 19b | D;V | Give an overall interpretation of the results, considering objectives, limitations, results from similar studies, and other relevant evidence. | 22 |
| Implications | 20 | D;V | Discuss the potential clinical use of the model and implications for future research. | 22-23 |
| **Other information** | | | | |
| Supplementary information | 21 | D;V | Provide information about the availability of supplementary resources, such as study protocol, Web calculator, and data sets. | Supplementary data |
| Funding | 22 | D;V | Give the source of funding and the role of the funders for the present study. | 2 |
